# Development and Validation of a Point-of-Care Triage Scorecard to Enhance Tuberculosis Case Detection During Active Community Screening in Yogyakarta, Indonesia

**DOI:** 10.64898/2026.08.27.26361569

**Authors:** Dani Catrianiningsih, Felisia, Bintari Dwihardiani, Albarissa Shobry Abdalla, Sepsiana Puspitasari, Rina Triasih

## Abstract

In primary healthcare centers lacking advanced imaging, community-based active tuberculosis (TB) case finding often relies on basic symptom screening. This approach often misses cases and leads to the inefficient allocation of rapid molecular testing (RMT). We aimed to develop and internally validate a simple clinical triage scorecard to improve TB detection and guide RMT use in resource-constrained settings. We conducted a retrospective cross-sectional study of 15,137 adults (≥18 years) evaluated within the Zero TB Yogyakarta program (2020-2025). Participants with complete clinical assessments and confirmatory GeneXpert results were included. Using multivariable logistic regression, we identified independent clinical predictors, which were subsequently transformed into an integer-based point scorecard. Model performance was evaluated via discrimination and calibration, utilizing bootstrap resampling (1,000 iterations) for internal validation. Among the 15,137 participants, 251 (1.7%) were GeneXpert-positive. The final multivariable model identified eight independent predictors: age, male sex, body mass index, prolonged cough, hemoptysis, unexplained weight loss, TB contact history, and diabetes mellitus. The model demonstrated strong predictive accuracy, with an optimism-adjusted AUROC of 0.836 and good calibration. When translated to the integer scorecard and compared directly to standard national symptom screening, the scorecard performed significantly better (AUROC 0.81 vs. 0.73; p<0.001). At a high sensitivity cut off score of ≥ 0, the tool achieved 93.63% sensitivity and 41.33% specificity. This point-of-care clinical scorecard provides higher diagnostic accuracy than standard symptom screening algorithms. By offering flexible operational thresholds, it empowers local health programs to dynamically balance the urgency of case detection with available diagnostic capacity, optimizing GeneXpert allocation where advanced radiological imaging is unavailable.

## Introduction

Tuberculosis (TB) remains a major global health crisis, with an estimated 10.7 million cases reported in 2024. Indonesia carries the second-highest TB burden globally, with an estimated incidence of 1,090,000 cases [1]. Halting the epidemic requires interrupting transmission chains through early detection and expanded active case finding (ACF) [2]. In response, a primary focus of the Indonesian Ministry of Health (MoH) is expanding ACF programs and improving diagnostic capabilities through scaled-up TB screening using portable X-rays (CXR) and rapid diagnostic tests [3]. In 2021, rapid molecular testing (e.g., GeneXpert) officially replaced acid-fast bacilli (AFB) microscopy as the primary diagnostic gold standard in Indonesia [4]. However, molecular diagnostic tests are resource-intensive and expensive, making it unfeasible to universally test every suspected patient and imposing a significant financial burden on local healthcare systems in resource-limited settings [5–7].

To optimize resource allocation, the World Health Organization (WHO) recommends systematic screening algorithms utilizing tools such as standard symptom screening, AI-augmented chest radiography (AI-CXR), and C-reactive protein (CRP) [8]. Standard symptom screening exhibits low-to-moderate sensitivity (30–70%) but moderate-to-high specificity (65–90%). Conversely, AI-CXR achieves high sensitivity (85–95%), though its specificity varies significantly depending on the operational threshold applied [8,9]. Selecting an appropriate triage algorithm requires navigating the operational trade-off between maximizing sensitivity for case detection and maximizing specificity to conserve diagnostic resources.

While expanding ACF is a national priority, uneven diagnostic capacity across the archipelago complicates implementation because advanced testing is concentrated in selected facilities and regions. Although the AI-CXR screening shows strong promise for improving TB detection, its rollout still faces major structural barriers, including high costs, infrastructure and maintenance requirements, and the need for reliable electricity and supporting health-system capacity [10,11] . Consequently, the Indonesian National Tuberculosis Program (NTP) mandates parallel screening (AI-CXR combined with symptom screening) where available, but relies solely on symptom screening for puskesmas (primary healthcare centers) lacking radiological facilities. Currently, access to CXR services at these centers remains severely limited; none of the study sites offer CXR services, and nationwide availability of CXR services at primary care facilities is below 30%. Because traditional symptom-based definitions of presumptive TB frequently miss cases, maximizing alternative, practical clinical measures is critical. Incorporating demographic and extended clinical predictors into a unified assessment has the potential to improve triage accuracy [9].

The absence of a highly discriminative clinical tool capable of operating entirely at the point of care without radiological support remains a critical gap in current clinical practice. To address these limitations, primary healthcare facilities require an optimized scorecard that bridges the gap between basic symptom screening and advanced imaging. A continuous scoring system empowers local health programs to flexibly adjust diagnostic thresholds, safely navigating the diagnostic paradox based on their specific epidemiological context and laboratory capacity. Therefore, this study aims to address the following question: What is the diagnostic performance and operational utility of a novel, internally validated clinical scorecard for tuberculosis triage when compared to standard symptom screening and AI-augmented chest radiography?

## Materials and Method

### Study Design and Population

This study and the subsequent reporting of the multivariable prediction model were conducted in accordance with the TRIPOD+AI (Transparent Reporting of a multivariable prediction model of Individual Prognosis Or Diagnosis, including AI) guidelines [12]. This study utilized retrospective data collected through the Zero TB Yogyakarta active case finding program, which conducted community-based screening between February 2020 and September 2025. While these underlying field screening activities were conducted in accordance with the standard operational protocol of the broader project, a separate protocol was not prepared, and this retrospective model development study was not registered in a public database. The screening program operated in an epidemiological setting where the baseline regional TB incidence in the Special Region of Yogyakarta ranges from 250 to 300 per 100,000 population [13], compared to a national incidence rate of 387 per 100,000. During the program, all ACF participants underwent parallel symptom and chest radiograph (CXR) screening. In accordance with the Indonesian National Tuberculosis Program (NTP) guidelines, participants were classified as having presumptive TB if they presented with specific clinical criteria: a cough lasting at least two weeks, haemoptysis, night sweats, unexplained weight loss, any symptom if living with HIV (PLHIV), or a cough of any duration accompanied by at least one other classic TB symptom. Following the conclusion of these field activities, the programmatic dataset was accessed on June 23, 2026, for this research. During data curation and analysis, the authors had access to information that could identify individual participants. To safeguard participant privacy, all identifiable data were stored on secure, password-protected institutional servers, with access strictly restricted to the core research team.

The criteria for a positive CXR screen evolved during the study period. Prior to September 2021, CXR was deemed positive based on manual interpretation by ACF physicians. From September 2021 onward, CXR screening incorporated computer-aided detection (CAD; qXR.ai version 4.1), and a positive screen was defined by a CAD score above a 0.5 threshold. Any participant who screened positive via either the NTP symptom criteria or the CXR evaluation (manual or CAD-interpreted) underwent confirmatory sputum testing. The primary predicted outcome for this analysis was a bacteriologically confirmed positive GeneXpert result.

All clinical, demographic, and anthropometric predictors were evaluated at the point of care during the initial active case finding encounter. Trained ACF staff administered a standardized, structured screening questionnaire to capture self-reported demographic characteristics, symptom profiles, and medical history. Anthropometric measurements were obtained objectively on-site. Because all clinical and demographic assessments were completed and recorded before sputum samples were processed for molecular testing, the assessment of all predictor variables was naturally blinded to the final GeneXpert reference outcome, effectively preventing interpretation bias or data leakage. The specific categories and units for all evaluated predictors are detailed in Table 1.

**Table 1.**
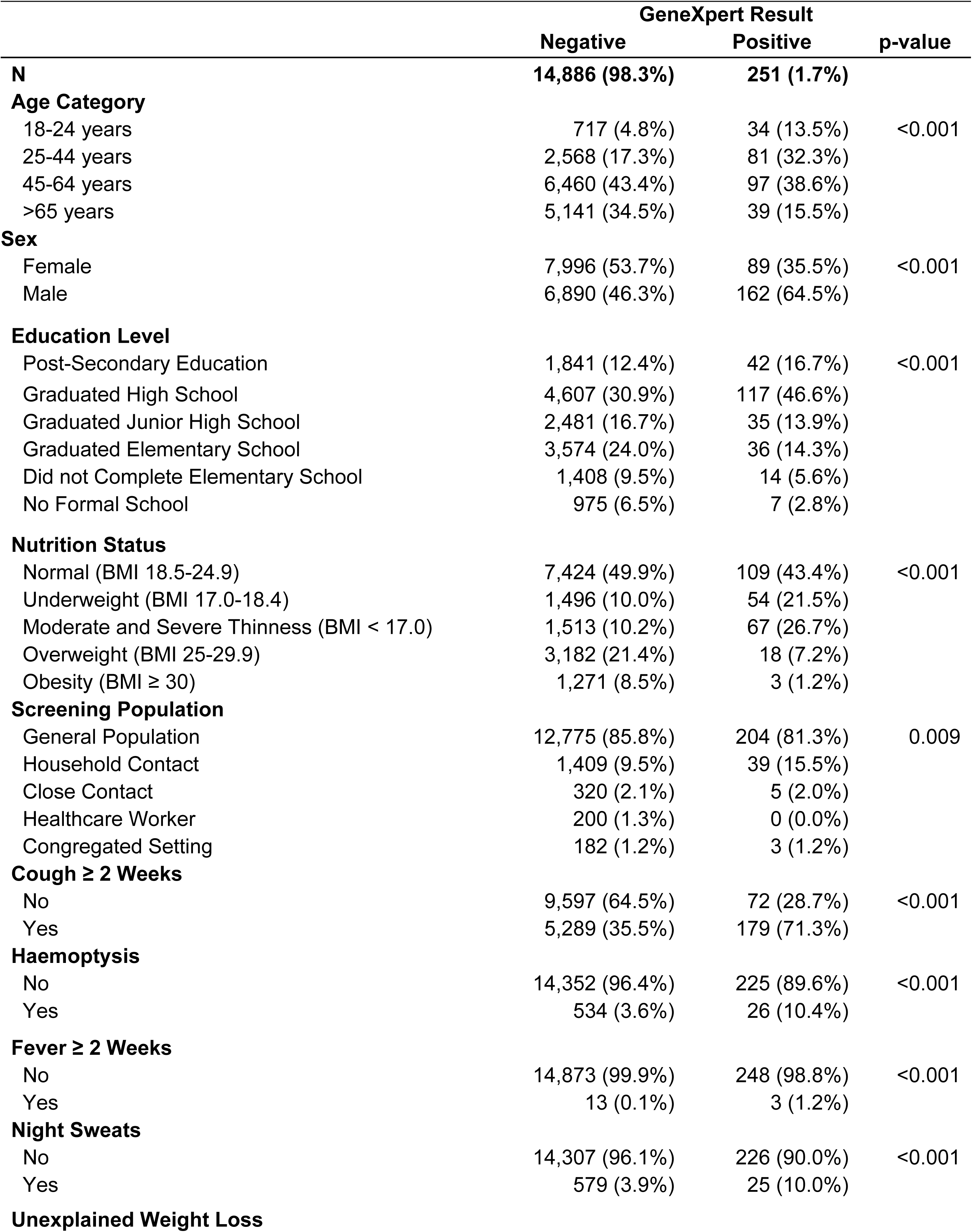
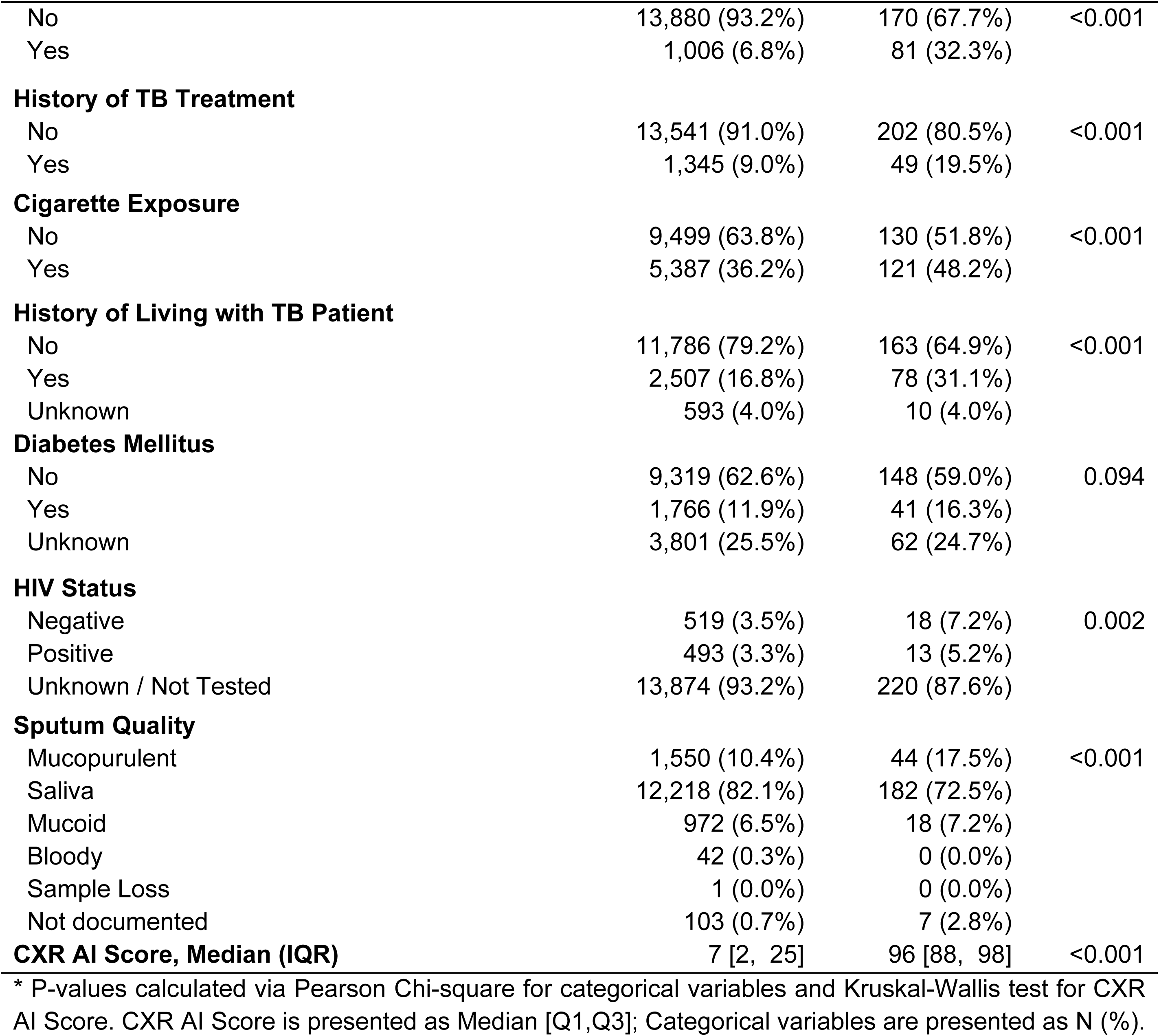
Baseline characteristics of the model development cohort, stratified by GeneXpert positivity.

### Data Management

Data was collected using Research Electronic Data Capture (REDCap) software hosted at Universitas Gadjah Mada. Data management procedures included consistency checks, the removal of duplicate records, and the assessment of missing values. All statistical analyses were subsequently performed using Stata version 19 (StataCorp, College Station, TX, USA).

### Predictor Selection and Sample Size

Standard methodological guidelines for the development of clinical prediction models recommend a minimum of 10 to 15 outcome events per candidate predictor parameter (events per variable, or EPV) to ensure model stability and minimize the risk of overfitting [14]. To prevent model overfitting given the limited number of primary outcome events (n = 251 GeneXpert positive cases), candidate predictors were pre-selected prior to statistical modeling. Initial predictor selection was guided by a review of literature to identify established clinical, demographic, and socioeconomic risk factors for tuberculosis. These evidence-based risk factors were then cross-referenced with the variables routinely collected within the Zero TB Yogyakarta ACF dataset. Through this process, 16 theoretical candidate variables were selected for the initial univariable screening phase. Once multi-level categorical variables were expanded, this accounted for a total of 25 distinct parameters, ensuring the model remained both clinically relevant and mathematically stable within the constraints of the available sample size.

Consequently, among the 15,137 eligible participants with complete records, the 251 confirmed TB cases provided an actual baseline EPV of 10.04 for the 25 initial parameters. Furthermore, following univariable screening, the 9 parameters that advanced to the final multivariable model yielded a substantially higher EPV of 27.89. This ratio ensures strong mathematical stability, precise estimation of the adjusted odds ratios, and adequate statistical power for the subsequent internal bootstrap validation protocols.

### Statistical Analysis and Model Development

The scorecard was developed using a cohort of participants aged ≥ 18 years who had complete clinical and demographic records and valid GeneXpert test results. Inclusion was not restricted by the availability of CXR results. Individuals undergoing TB treatment at the time of screening were excluded, yielding a final analytical dataset of 15,137 participant records. To maximize statistical power and ensure model stability given the limited number of primary outcome events, this entire cohort was utilized for model development. Consequently, the dataset was not partitioned into separate training and testing subsets; instead, internal validation and performance evaluation were conducted simultaneously using bootstrap resampling techniques.

To facilitate the translation of the multivariable model into a simple, integer-based clinical scorecard, continuous variables were categorized prior to analysis. Age was grouped into four distinct cohorts (18–24, 25–44, 45–64, and ≥ 65 years), and Body Mass Index (BMI) was stratified according to standard nutritional statuses. Furthermore, to maintain model stability and achieve minimum Events Per Variable (EPV) constraints, several categorical predictors were systematically collapsed. Participants identified within the screening population as "household contacts" or "close contacts" were merged into a single "Household or Close TB Contact" variable. For treatment history, individuals who had previously completed TB treatment and those who had defaulted were combined into a unified "History of TB Treatment" predictor (individuals currently undergoing active TB treatment were excluded from the study cohort entirely). For smoking status, past and current smokers were combined into a single "Cigarette Exposure" category. Finally, for both Diabetes Mellitus and HIV status, "negative" and "unknown/not tested" responses were collapsed into a single reference category. This pragmatic grouping reflects the operational realities of community-based active case finding, where baseline point-of-care testing for these specific comorbidities is frequently unavailable.

Baseline clinical and demographic characteristics were summarized using frequencies and percentages for categorical variables and means with standard deviations or medians with interquartile ranges (IQRs) for continuous variables. Because the primary predicted outcome (GeneXpert positivity) is binary, multivariable logistic regression was selected as the optimal modeling approach. This method was specifically chosen because its resulting adjusted β coefficients can be reliably translated into a simple, additive point system, aligning directly with the operational need for a non-digital triage tool. To identify independent predictors of GeneXpert positivity, univariable logistic regression was performed. Candidate predictors with a p < 0.20 or documented clinical significance (e.g., PLHIV status) were advanced to multivariable modeling. For multi-category variables, global p-values were evaluated using the Wald test. In the multivariable logistic regression model, a purposive manual backward step-down elimination method sequentially removed non-significant variables (p ≥ 0.05). Confounding was assessed at each reduction step; variables were retained if their removal altered the odds ratio (OR) of any remaining predictor by > 20%. To ensure model stability, multicollinearity among the final predictors was evaluated using the variance inflation factor (VIF), with values < 5.0 considered acceptable.

To translate the final multivariable regression model into a practical, point-of-care triage scorecard, an integer-based point system was derived. Adjusted β coefficients were scaled by dividing each β by the coefficient of the smallest significant binary predictor, and the quotient was rounded to the nearest whole integer.

Model performance was evaluated across three primary domains: discrimination, calibration, and clinical utility. Discrimination was quantified using the area under the receiver operating characteristic curve (AUROC), with internal validation and overfitting correction performed via Harrell’s bootstrap resampling method (1,000 iterations) to derive an optimism-adjusted AUROC. Calibration was assessed statistically using the Hosmer-Lemeshow goodness-of-fit test and evaluated visually via a calibration plot (S1 Fig). The final output of the prediction model is a continuous, integer-based risk score. To assess clinical utility and establish practical classifications, diagnostic metrics including sensitivity, specificity, and the Youden Index were calculated across all integer cut-offs. Rather than imposing a single, rigid binary classification, this approach allowed for the establishment of flexible, operational risk tiers tailored to local epidemiological contexts and available laboratory capacities. To compare the diagnostic performance of the derived scorecard against existing triage modalities (the AI-CXR strategy and standard NTP symptom screening), differences between the respective AUROCs were evaluated for statistical significance utilizing DeLong’s non-parametric test. Finally, to evaluate model fairness and ensure unbiased performance across diverse patient populations, subgroup analyses were conducted. Discriminatory performance (AUROC with 95% confidence intervals) was calculated separately for key sociodemographic and clinical categories, including sex, age, nutritional status, and HIV status.

### Model Triage Comparison

To assess the diagnostic utility of the clinical scorecard relative to the single AI-CXR and the parallel algorithm (AI-CXR plus symptoms), we performed a direct comparative analysis. Although model derivation and internal validation utilized the entire study population (n=15,137) to ensure maximum stability, the comparative performance evaluation was restricted to a uniform dataset containing valid results across all screening modalities. Consequently, this analysis was conducted on a complete-case subcohort of 10,732 individuals with concurrent clinical and demographic assessments, GeneXpert test results, and AI-CXR scores. Missing radiological data for the excluded subset resulted entirely from programmatic and operational barriers encountered during fieldwork. These challenges involved temporary technical issues with mobile X-ray units and the staggered implementation of the AI-CXR software during the ACF project’s inception. As a result, these data were classified as missing completely at random (MCAR), minimizing the likelihood of selection bias within the comparison. Baseline characteristics for this analytic subcohort are presented in S1 Table to confirm population comparability.

### Ethics Approval

The study protocol, including the retrospective analysis of programmatic active case finding (ACF) data, was reviewed and formally approved by the Medical and Health Research Ethics Committee (MHREC) of Universitas Gadjah Mada (UGM), Yogyakarta, Indonesia. Written informed consent was obtained from all participants prior to their initial clinical screening and inclusion in the Zero TB Yogyakarta ACF program. For this retrospective study, the MHREC approved the research team’s access to identifiable participant data required for secondary analysis. Strict data security protocols were enforced throughout the study to maintain patient confidentiality, and all identifiable information was stripped from the analytical dataset prior to the dissemination or publication of these findings.

### Patient and Public Involvement

While patients and the public were not directly involved in the retrospective data analysis or the statistical design of the prediction model, there was ongoing engagement with local stakeholders, puskesmas, community health workers, and the general public throughout the underlying Active Case Finding (ACF) program.

## Result

### Participant Characteristics

The flow of participants through the study, including exclusions and final cohort sizes, is detailed in Fig 1.

**Fig 1.**
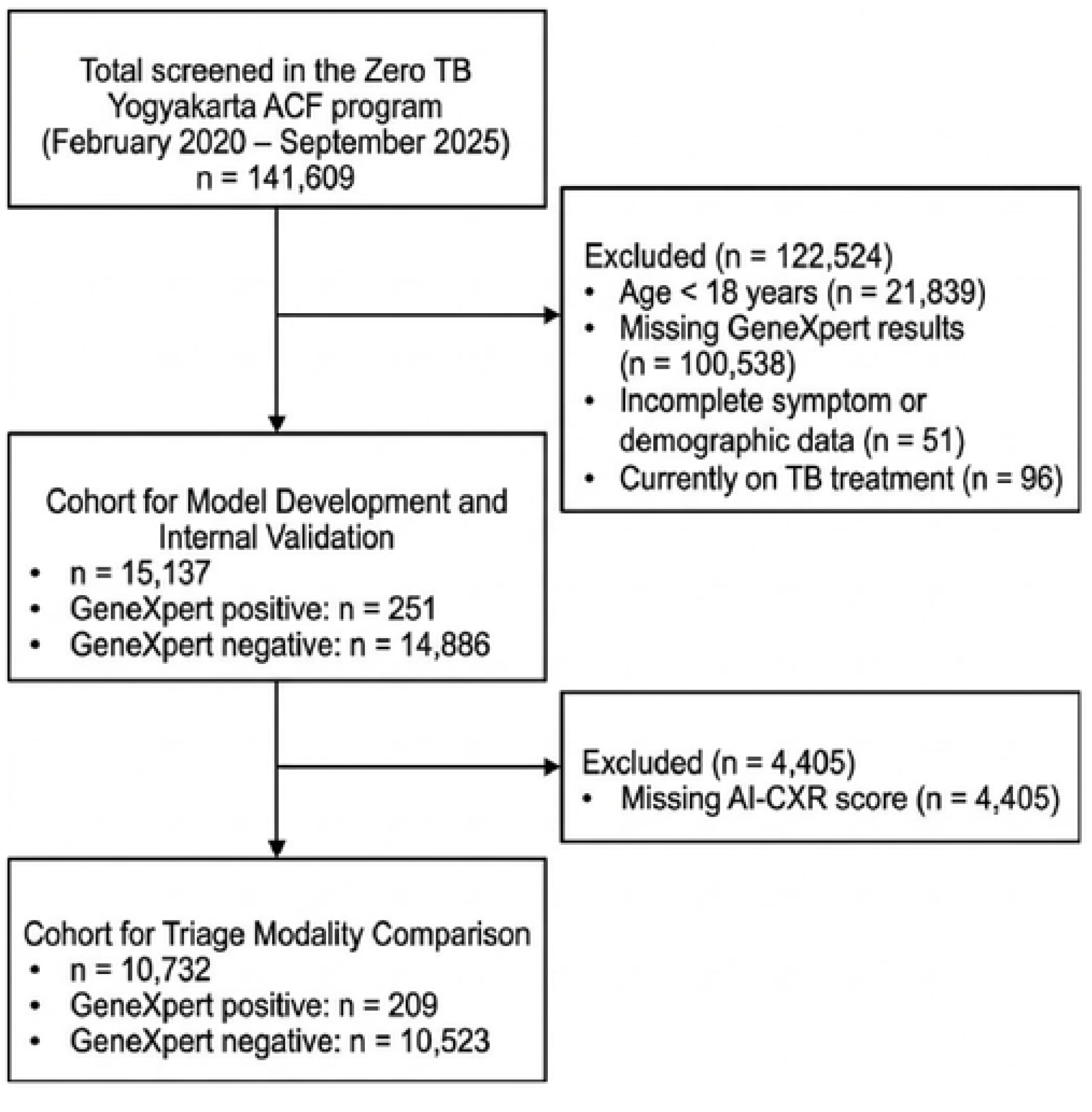
Participant flow diagram detailing the derivation of the model development and triage comparison cohorts.

A total of 15,137 participants were included in the final analysis, of whom 251 (1.7%) had positive GeneXpert MTB results. Participant characteristics according to GeneXpert results are summarized in Table 1.

### Predictive Model Development and Collinearity Assessment

Following univariable screening and manual backward step-down elimination, the final multivariable model retained nine clinical and demographic predictors (Table 2). Model stability was confirmed, with all independent clinical features maintaining individual VIF values below 2.0, aside from the expected structural multicollinearity within the multicategory age variable. In the adjusted analysis, male sex, undernutrition, prolonged cough, hemoptysis, unexplained weight loss, close TB contact, and diabetes mellitus emerged as independent risk factors for GeneXpert positivity. Conversely, older age and overweight or obese BMI demonstrated protective associations. Notably, controlling for confounding covariates markedly accentuated the adjusted odds ratios for both close TB contact and diabetes mellitus, whereas HIV status. Remained non-significant in both the unadjusted and adjusted model.

**Table 2.**
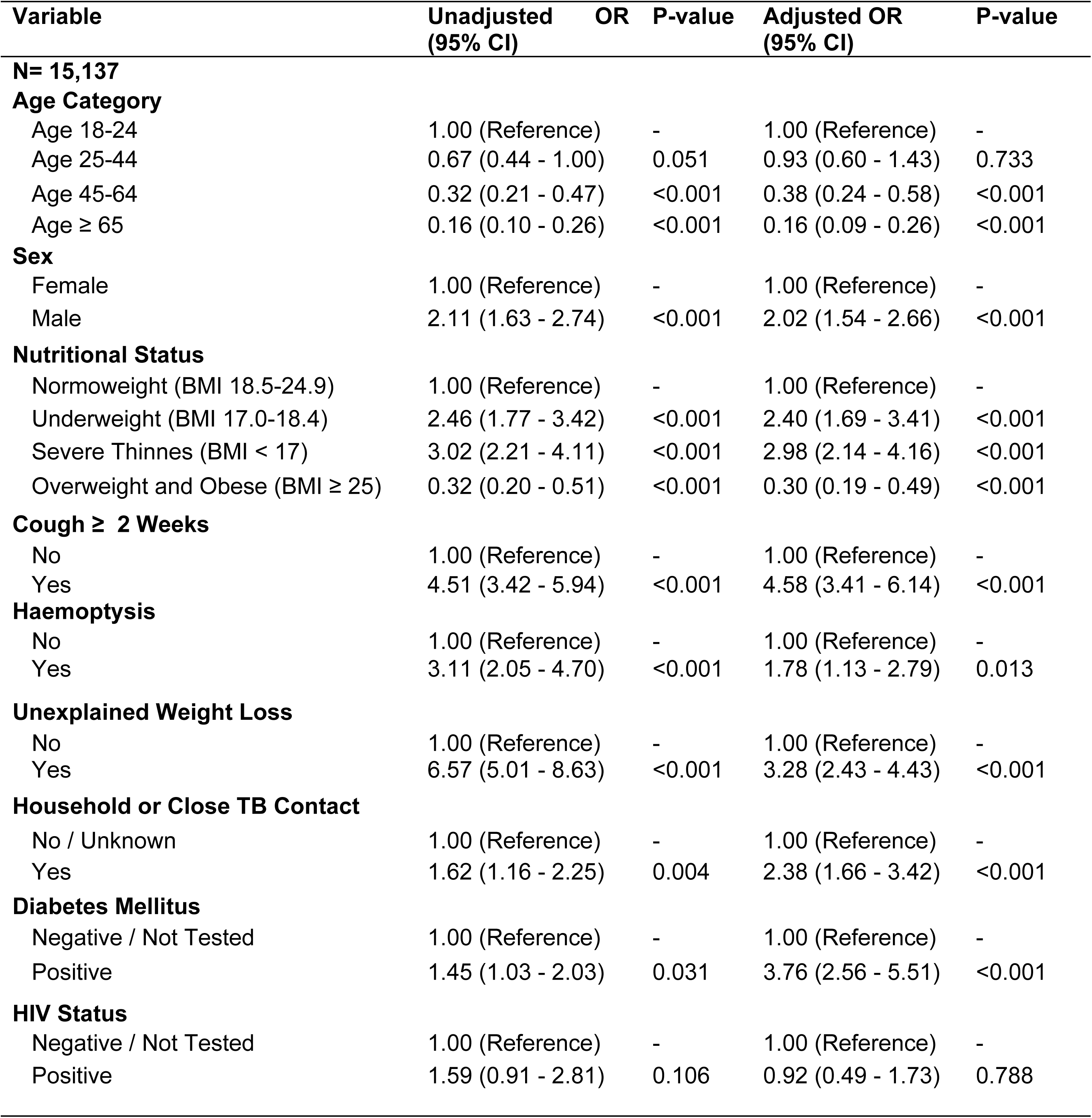
Univariable and Multivariable Logistic Regression Predictors of GeneXpert Positivity.

### Translation into the Additive Scoring System

To translate the final multivariable regression model into a practical screening tool, the adjusted β coefficients were scaled relative to the smallest significant positive binary predictor (haemoptysis, β = 0.57) and rounded to the nearest integer to assign point values (Table 3). Within this additive scoring system, a prolonged cough carries the highest individual risk weight. Conversely, advancing age cohorts and an overweight nutritional status contribute negative points to the total score, reflecting their independent protective associations against GeneXpert positivity.

**Table 3.** Point Allocation Matrix for the TB Scorecard.

| Predictor Variable | Category / Value | Assigned Points |
| --- | --- | --- |
| **Demographics** |  |  |
| Sex | Female | 0 (Reference) |
|  | Male | +1 |
| Age Category | 18–24 years | 0 (Reference) |
|  | 25–44 years | 0 |
|  | 45–64 years | -2 |
|  | ≥65 years | -3 |
| **Clinical Symptoms** |  |  |
| Cough ≥ 2 Weeks | No / Yes | 0 (Ref) / +3 |
| Haemoptysis | No / Yes | 0 (Ref) / +1 |
| Unexplained Weight Loss | No / Yes | 0 (Ref) / +2 |
| **Nutritional Status** |  |  |
|  | Normoweight | 0 (Reference) |
|  | Underweight | +2 |
|  | Overweight | -2 |
| **Risk Factors** |  |  |
| Household or Close TB Contact | No or Unknown / Yes | 0 (Ref) / +2 |
| Diabetes Mellitus | No or Unknown / Yes | 0 (Ref) / +2 |

### Diagnostic Performance Across Score Thresholds

To evaluate the clinical application of the scorecard, diagnostic performance metrics were calculated across all possible score thresholds (S2 Table). At a threshold of ≥ 0, the scorecard achieved a sensitivity of 93.63% and a specificity of 41.33%. Increasing the threshold to ≥ 1 resulted in a specificity of 57.83% and a sensitivity of 87.25%. Although a threshold of ≥ 2 mathematically maximized the Youden Index (47.90), this cut point yielded a lower sensitivity of 76.49%.

### Comparative Performance of Triage Strategies

The discriminatory power of the developed scorecard was evaluated against standard NTP symptom screening and the AI-augmented chest radiography (AI-CXR) strategy using receiver operating characteristic (ROC) curve analysis (Figure 2). The clinical scorecard achieved an area under the ROC curve (AUROC) of 0.81 (95% CI: 0.79–0.84). This represented a statistically significant improvement in discriminatory ability compared to the standard NTP symptom screening, which yielded an AUROC of 0.73 (95% CI: 0.71–0.75; p < 0.001). The AI-CXR strategy demonstrated the highest overall discrimination among the three modalities, achieving an AUROC of 0.89 (95% CI: 0.86– 0.91; overall p < 0.001).

**Fig 2.**
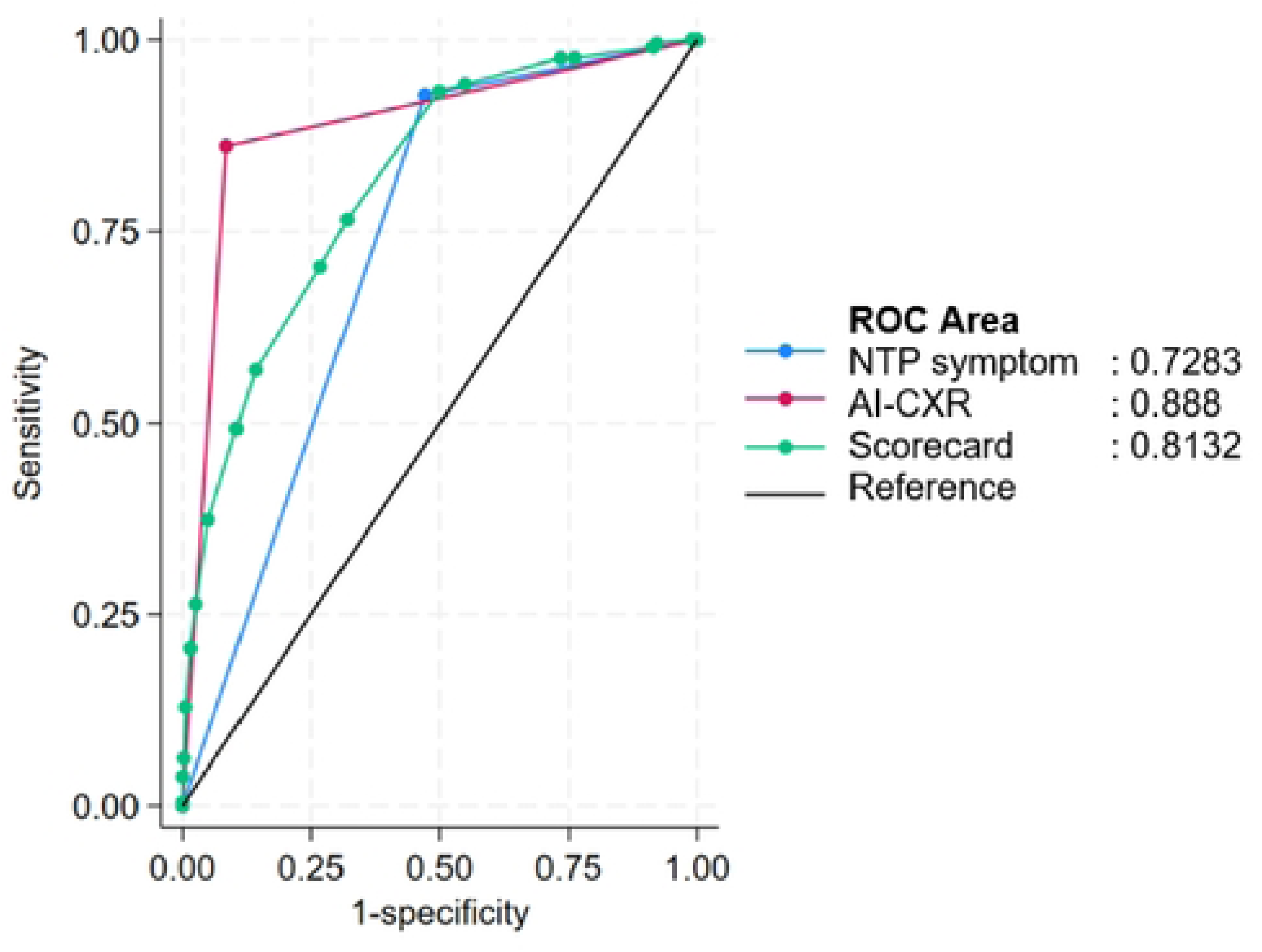
Receiver operating characteristic (ROC) curves comparing the diagnostic performance of AI-CXR, Scorecard, and NTP symptom screening. The areas under the receiver operating characteristic curve (AUROC) were 0.888 (95% Cl: 0.864–0.912) for AI-CXR, 0.813 (95% Cl: 0.785–0.841) for the Clinical Scorecard, and 0.728 (95% Cl: 0.710–0.746) for NTP symptom screening.

Subgroup analysis demonstrated consistent model discrimination across key demographic and clinical categories, with AUROCs ranging from 0.707 to 0.859. Detailed performance metrics, including 95% confidence intervals stratified by sex, age, nutritional status, and HIV status, are provided in S3 Table.

### Model Performance Evaluation and Internal Validation

Within the evaluation cohort (n = 15,137), the model achieved an area under the receiver operating characteristic curve (AUROC) of 0.84 (95% CI: 0.82–0.87). The Hosmer-Lemeshow goodness-of-fit test was non-significant (***χ***^2^ = 5.70, *p* = 0.68). Following Harrell’s optimism correction via bootstrapping with 1,000 replications, the model exhibited an average optimism of 0.0045, resulting in a final optimism-adjusted AUROC of 0.836.

## Discussion

In this retrospective analysis of 15,137 adults undergoing community based active case finding (ACF), we developed and internally validated a novel, eight-variable clinical triage scorecard to optimize tuberculosis case detection. The derived scorecard significantly outperformed standard National TB Program (NTP) symptom screening, capturing a broader risk profile that traditional, purely symptom-based approaches often miss. Furthermore, while AI-augmented chest radiography (AI-CXR) maintained the highest overall discrimination, the scorecard emerged as a highly competitive and robust non-radiological alternative. By mathematically optimizing readily available demographic, anthropometric, and clinical predictors, this tool bridges the critical performance gap between basic clinical screening and advanced imaging.

### Clinical Utility as a Non-Radiological Alternative

The clinical scorecard demonstrated a statistically significant improvement in discriminatory power (AUROC 0.81) compared to the standard NTP symptom-based screening guidelines (AUROC 0.73; *p* < 0.001). Traditional symptom-based screening algorithms frequently miss patients with asymptomatic or atypical presentations [[15]. By mathematically optimizing a comprehensive set of demographic and clinical predictors, the scorecard captures a broader risk profile, significantly reducing the number of missed cases without the need for additional hardware. Incorporating these demographic variables alongside clinical symptoms is particularly valuable because tuberculosis operates as a social pathology; an individual’s risk is inextricably linked to social, biological, and pathological characteristics—such as age, sex, and underlying comorbidities like malnutrition and diabetes—that traditional, symptom-only screens frequently fail to capture [9]. This magnitude of improvement aligns with previous international efforts to convert symptom screening into graded risk estimates. A scoring system derived from a large screening trial in South Africa and validated in Zambia similarly outperformed both prolonged cough alone (AUC 0.58) and the presence of any TB symptom (AUC 0.60), although its absolute discrimination remained lower (0.69 and 0.66, respectively) than the performance achieved by our model.

Furthermore, when evaluated alongside existing triage modalities, the clinical scorecard provides a highly competitive, non-radiological alternative. The AI-CXR strategy yielded the highest overall discrimination (AUROC 0.89), a finding consistent with independent multi-country evaluations reporting AUCs between 0.85 and 0.92 for AI-CXR against microbiological reference standards in community-based ACF and migrant screening [16,17]. However, radiological infrastructure is not universally available. For puskesmas operating in resource-constrained settings that lack access to X-ray equipment, implementing AI-CXR remains logistically challenging. While AI-augmented imaging remains the ideal standard for triage, this scorecard bridges the critical performance gap between basic symptom screening and advanced imaging entirely at the point of care, serving as an immediate solution for optimizing GeneXpert allocation. Subgroup analysis demonstrated consistent discrimination across key demographic and clinical categories, with AUROCs ranging from 0.707 to 0.859. Detailed performance metrics, including 95% confidence intervals stratified by sex, age, nutritional status, and HIV status, are provided in S3 Table.

### Epidemiological Alignment of Scorecard Predictors

The variable weightings derived for this scorecard align strongly with established epidemiological evidence. Key clinical and demographic risk factors, including a prolonged cough (+3 points), unexplained weight loss (+2 points), close TB contact (+2 points), undernutrition (+2 points) and a history of diabetes mellitus (+2 points), emerged as the primary drivers of the predictive model. The substantive weight assigned to close TB contact is well supported by systematic reviews demonstrating that the prevalence of active TB among household contacts in low- and middle-income settings is approximately 3.1%, a rate several-fold higher than the background community prevalence [18]

Interestingly, the model also captured the protective epidemiological effects of advancing age and higher nutritional status within this specific cohort. The scorecard applies to a point deduction for individuals who are overweight or obese (-2 points), which is consistent with extensive cohort studies and meta-analyses demonstrating a strong inverse relationship between a higher body mass index (BMI) and the risk of developing active tuberculosis [19,20]. Furthermore, advanced age led to greater point deductions, resulting in a -3-point reduction for individuals aged 65 years and older. From a programmatic perspective, this age dynamic likely reflects the specific nature of community-based active case finding (ACF). The higher odds of GeneXpert positivity in younger demographics may be driven by the increased community exposure, occupational clustering, and mobility among the productive-age population, particularly males, within the screened communities.

Notably, HIV status did not demonstrate a statistically significant association with GeneXpert positivity in the multivariable analysis. People living with HIV are estimated to be 12 to 16 times more likely to develop TB disease than the general population, with the risk rising steeply as CD4 counts decline [[21,22]. Therefore, the absence of an association in our model represents a limitation of the dataset rather than a substantive clinical finding. This lack of significance is likely attributable to the specific epidemiological profile of the Yogyakarta region, which features a considerably lower baseline HIV prevalence compared to other high-burden TB settings. Additionally, variables such as HIV and diabetes mellitus status were obtained via self-reported anamnesis during the high-throughput ACF workflow, where approximately 100 participants were screened daily, rather than through objective point-of-care testing. Consequently, the model’s reliance on unverified self-reporting for these heavily stigmatized or underdiagnosed comorbidities may have resulted in underreporting, thereby dampening their statistical impact on the final scorecard.

### Navigating the Diagnostic Paradox via Flexible Thresholds

For practical applications, this scorecard offers flexibility to adapt to local diagnostic capacities. This adaptability is programmatically meaningful because molecular testing capacity, rather than screening coverage, is frequently the primary binding constraint in high-burden settings. In fact, universal door-to-door universal sputum screening has been shown to be substantially more costly per case than targeted approaches [23]. By providing a continuous, integer-based scoring system, this tool allows local health policymakers to dynamically adjust their triage strategy, directly addressing the "diagnostic paradox" inherent in community-based active case finding.

Because the core objective of a TB triage tool is to maximize case detection without overwhelming limited laboratory resources, selecting a practical cut point requires navigating the strict trade-off between diagnostic sensitivity and operational specificity. As demonstrated in our results, applying a cutoff score of ≥ 0 strongly prioritizes case finding, capturing most true positive cases with an exceptional sensitivity of 93.63%.

However, this aggressive threshold yields a low specificity of 41.33%, which risks massively increasing the downstream volume of required GeneXpert assays. Increasing the threshold to ≥ 1 offers a highly pragmatic and balanced alternative; it significantly reduces operational testing costs by improving specificity to 57.83%, while still maintaining a robust diagnostic sensitivity of 87.25%. Conversely, while a threshold of ≥ 2 mathematically maximizes the Youden Index, the resulting drop in sensitivity to 76.49% renders this cut point clinically suboptimal for a frontline triage strategy, where minimizing false negatives remains paramount. Ultimately, the provision of flexible operational thresholds empowers local health programs to make evidence-based implementation decisions. It allows clinics to carefully balance the epidemiological urgency of case detection with their specific budgetary realities and available rapid molecular testing (RMT) capacity. However, for practical implementation, the tool requires the complete collection of all eight variables to ensure predictive accuracy; if objective measurements cannot be obtained, or if symptom history is highly unreliable, the scorecard should not be used as the sole triage determinant. Because it relies on simple integer addition, the scorecard requires no specialized computational or radiological expertise; it is designed to be easily administered by frontline health workers with basic clinical training to measure anthropometrics and elicit symptom history.

### Methodological Strengths and Study Limitations

Our model is also methodologically robust, having been trained and evaluated using a substantial screening cohort (n = 15,137). The derived scorecard demonstrated strong discriminatory power (AUROC 0.841) and excellent calibration, as evidenced by a non-significant Hosmer-Lemeshow goodness-of-fit test. Furthermore, the internal validation utilizing Harrell’s bootstrap resampling yielded a final optimism-adjusted AUROC of 0.836. Because this adjusted value is nearly identical to the original unadjusted AUROC, it strongly confirms the stability and reliability of the model’s predictive accuracy.

Despite these promising results, several critical limitations must be addressed before widespread implementation. First, while the overall cohort was large, the positive case count was relatively low (251/15,137), reflecting the specific epidemiological context of the Yogyakarta province, which possesses a considerably lower TB prevalence compared to other regions in Indonesia. Second, because the scorecard was developed using individuals presenting an active case finding (ACF) setting, there is a risk of referral bias. Half of the cohort met the NTP symptom criteria for presumptive TB, likely because many participants were actively referred by local health cadres due to perceived high risk. Consequently, the scorecard’s applicability as a mass screening tool for an entirely unselected, asymptomatic general population remains uncertain and requires further investigation. While our internal validation is statistically rigorous, the essential next step is external validation in diverse geographic and demographic contexts to ensure true generalizability.

Finally, this study focused strictly on diagnostic accuracy and did not measure direct operational impacts in the field, such as cost-effectiveness, the number needed to test (NNT), or the turnaround time for diagnosis. Moreover, the model was developed using GeneXpert positivity as the sole dependent variable. While the scorecard successfully optimizes the allocation of this specific assay, its reliance on a single rapid molecular testing (RMT) platform limits the immediate applicability of the findings to other diagnostic infrastructures. Future real-world implementation studies and randomized controlled trials are required to confirm that the improved discrimination of this scorecard translates to increased case notification rates across various RMT platforms without overburdening local healthcare systems.

## Conclusion

The derived scorecard is a highly effective, non-radiological triage tool for community-based tuberculosis active case finding that significantly outperforms current symptom-based screening. By offering flexible diagnostic thresholds, it empowers health programs to balance case detection urgency with limited molecular testing capacity. While external validation is required, this tool provides an immediate, practical solution to optimize GeneXpert allocation in resource-constrained settings where advanced imaging is unavailable.

## Acknowledgement

The authors acknowledge the support and guidance provided by the Genomics and Science Dojo, the Genomics and Science Workshop, and its facilitators during manuscript preparation. The Genomics and Science Dojo and Workshop were conducted by the Summit Institute for Development, the GSI Lab, and the Oxford University Clinical Research Unit–Jakarta, with support from the British Embassy in Jakarta (FCDO Programme Name: Genomics and Science DOJO 3.0: Accelerating Scientific Outputs and Strategic Collaboration with Multisectoral Organisations; FCDO Project Number: 400422-412).

## Data Availability Statement

The datasets analyzed during the current study cannot be shared publicly due to ethical restrictions and the inclusion of sensitive, identifiable patient health information from the Active Case Finding (ACF) program. Data access is restricted by the Medical and Health Research Ethics Committee (MHREC) of Universitas Gadjah Mada to protect patient confidentiality.

However, the minimal data set underlying the findings of this study is available to peer reviewers and qualified researchers who meet the criteria for access to confidential data. Data access requests should be directed to the Data Management Committee of the Zero TB Yogyakarta, Center for Tropical Medicine, Universitas Gadjah Mada (contact via).

The Stata analytical code necessary to reproduce the statistical findings of this study is included with this published article as Supplementary Information (S4 Code).

## Supporting Information

**S1 Table.** Baseline characteristics of the triage evaluation cohort, stratified by GeneXpert positivity. (DOCX)

**S2 Table.** Score cutpoint diagnostics and operating characteristics. (DOCX)

**S3 Table.** Scorecard performance (AUROC) across key subgroups. (DOCX)

**S1 Fig.** Calibration plot of the multivariable clinical scorecard. (TIF)

